# mRNA COVID-19 vaccine effectiveness against SARS-CoV-2 infection in a prospective community cohort, rural Wisconsin, November 2020-December 2021

**DOI:** 10.1101/2021.12.14.21267809

**Authors:** Huong Q McLean, David L McClure, Jennifer P King, Jennifer K Meece, David Pattinson, Gabriele Neumann, Yoshihiro Kawaoka, Melissa A Rolfes, Edward A Belongia

**Affiliations:** Marshfield Clinic Research Institute, Marshfield, Wisconsin, United States; Department of Pathobiological Sciences, School of Veterinary Medicine, University of Wisconsin-Madison, Wisconsin, United States; Centers for Disease Control and Prevention, Atlanta, Georgia, United States

**Keywords:** SARS-CoV-2, COVID-19, vaccine effectiveness

## Abstract

Reduced COVID-19 vaccine effectiveness (VE) has been observed with increasing predominance of the Delta variant. In a prospective rural community cohort of 1265 participants, VE against symptomatic and asymptomatic SARS-CoV-2 infection was 56% for mRNA COVID-19 vaccines.

## Background

Multiple studies have demonstrated high effectiveness of coronavirus disease (COVID-19) vaccines in real world settings [1]. However, some studies have found reduced vaccine effectiveness (VE) against severe COVID-19 caused by acute respiratory syndrome coronavirus 2 (SARS-CoV-2) Delta (B.1.617.2) variant among persons who are immunocompromised, and against symptomatic infection at longer time since vaccination [1-6]. Furthermore, most evaluations of COVID-19 VE have focused on prevention of medically-attended SARS-CoV-2 infection or on effectiveness in high-risk populations, such as healthcare workers. We conducted active surveillance in a well-defined rural community cohort to estimate the effectiveness of messenger RNA (mRNA) COVID-19 vaccines against symptomatic and asymptomatic laboratory-confirmed SARS-CoV-2 infection.

## Methods

This analysis used data collected from an ongoing prospective community cohort study to assess SARS-CoV-2 infection in rural central Wisconsin, United States. Participants were enrolled November 2020-March 2021, and actively monitored weekly (up to 52 weeks) to ascertain symptoms and identify new SARS-CoV-2 infections.

### Study population

Participants were randomly sampled community-dwelling individuals living in the Marshfield Epidemiologic Study Area (central region), a 14 zip code region in central Wisconsin that includes Marshfield and surrounding area. The population is ∼53,000, and 89% receive most of their care from Marshfield Clinic Health System (MCHS) [7]. Further description of cohort eligibility criteria are in **Supplemental Methods**.

### Weekly illness surveillance

Each week, all participants reported the absence or presence of specific symptoms. Anterior nasal swabs were self-collected (or parent-collected) for each qualifying illness episode. In addition, approximately half of the cohort was assigned to collect swabs weekly for the first 26 weeks. Further details on assignment to symptomatic vs weekly nasal swab collection are provided in **Supplemental Methods**. A qualifying illness was a new illness onset in the past 7 days and at least one of the following symptoms: cough, fever or chills, sore throat, muscle or body aches, loss of smell or taste, shortness of breath, or diarrhea. Participants were instructed to also report any new respiratory symptoms by phone as soon as symptoms developed.

### Other

Participants had laboratory-confirmed SARS-CoV-2 infection if a specimen collected during surveillance was positive for SARS-CoV-2 by real-time reverse transcription polymerase chain reaction (rRT-PCR, ThermoFisher Combo Kit) or if they tested positive from a clinical PCR-based test at MCHS. Dates and results of clinical SARS-CoV-2 tests and vaccination data were extracted from MCHS electronic health records and obtained from self-report. Additional information regarding vaccinations, vaccine eligibility in Wisconsin, collection of demographic information and serum samples, and laboratory methods are described in **Supplemental Methods**.

### Analysis

Participant characteristics were compared across groups using Chi-square or Wilcoxon rank-sum tests. VE against laboratory-confirmed SARS-CoV-2 infection was estimated using Cox proportional hazards models with time-varying vaccination status, respiratory sample collection frequency, and age. Person-time at risk began at enrollment for persons aged ≥16 years or when participants aged 12-15 years became age-eligible for vaccination (May 13, 2021 or 12^th^ birthday after May 13, 2021), and ended December 7, 2021, date of positive SARS-CoV-2 infection, date of withdrawal from the study, or date of last weekly survey (study week 52), whichever occurred first. Unvaccinated person-time was defined as time before receipt of the first dose. Vaccinated person-time began ≥14 days after receipt of the second dose. Person-time from receipt of the first dose through 13 days after the second dose, and after receipt of vaccine off-label (before age-eligible or mixed-product series) was excluded, as were days following receipt of a third dose. In addition, person-time was excluded after receipt of Johnson & Johnson (Janssen) vaccine due to low uptake in the population.

VE for any mRNA vaccine and for each mRNA vaccine product was calculated as (1– hazard ratio) x 100%; the hazard ratio represented the ratio of SARS-CoV-2 infections in two-dose vaccinated to unvaccinated person-time. VE against symptomatic SARS-CoV-2 infection was estimated by excluding participants with infection with no reported symptoms during the 2 weeks before and after the positive test result (asymptomatic). VE against the Delta variant was estimated by restricting person-time at risk to the period after June 21, 2021, when >50% of sequenced viruses in Wisconsin were Delta. Sensitivity analyses excluded persons who self-reported or had serologic evidence of prior SARS-CoV-2 infection. Analyses were conducted using SAS (version 9.4; SAS institute).

Marshfield Clinic Research Institute (MCRI)’s Institutional Review Board reviewed and approved the study protocol. The Centers for Disease Control and Prevention (CDC) ceded research oversight to MCRI (45 C.F.R. part 46; 21 C.F.R. part 56).

## Results

Of 1518 cohort participants, 1266 (83%) were aged ≥12 years and included in this VE analysis. By the end of follow-up, almost half (48%) received 2 doses of Pfizer-BioNTech vaccine, 26% received 2 doses of Moderna vaccine, and 26% were unvaccinated. Older adults, females, Non-Hispanic White participants, those who received the 2020-2021 influenza vaccine, and those who work in healthcare were more likely to be vaccinated (**Supplemental Table**). Most (76%) vaccinated participants received a second dose in January-April 2021 (**Supplemental Figure**). Moderna recipients tended to be older, have a preexisting medical condition, have public insurance, and longer median time since receipt of the second dose (238 [interquartile range (IQR) 223-256] days vs 217 [IQR 182-247] days, Wilcoxon *P*<.001) compared with Pfizer-BioNTech recipients.

Between November 3, 2020 and December 7, 2021, 118 (9%) SARS-CoV-2 confirmed infections were documented during follow-up; 6 were asymptomatic (2 received Pfizer-BioNTech), 4 were previously infected (all unvaccinated) and 51 (43%) were vaccinated (29% received Pfizer-BioNTech, 14% received Moderna). Mean age of those infected was 47.7 years and 59% were female (**Supplemental Table**). Among unvaccinated participants with infection, 11 (16%) sought care, and 4 (6%) were hospitalized. Among vaccinated participants with infection, 4 (8%) sought care, and 1 (2%) was hospitalized. Median time from receipt of the second dose to infection was 215 (IQR 163-241) days.

VE of mRNA vaccines against laboratory-confirmed infection (symptomatic and asymptomatic) was 56% (95% confidence interval [CI] 31-71), 65% (95% CI 37-81) for Moderna, and 50% (95% CI 21-69) for Pfizer-BioNTech (**Figure**). VE estimates were similar against symptomatic infections, when prior infections were excluded, and when restricted to the period when Delta predominated (**Figure**).

**Figure 1.**
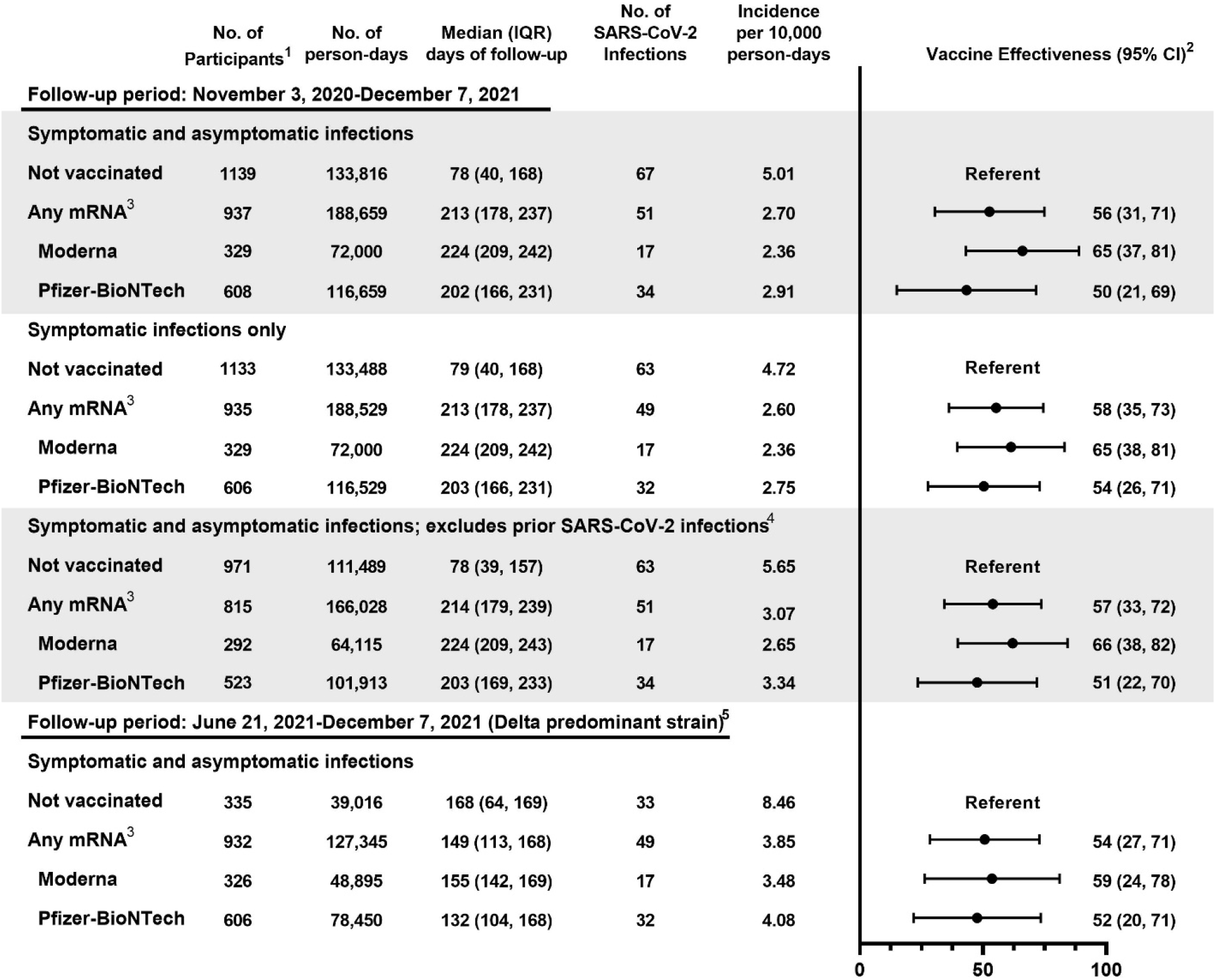
Effectiveness of COVID-19 vaccines against SARS-CoV-2 infection in a rural community, Wisconsin, November 3, 2020-December 7, 2021. ^1^Participants can contribute both unvaccinated and vaccinated time during the follow-up period. ^2^Estimated from Cox proportional hazards model with time-varying vaccination status, respiratory sample collection frequency, and age (modeled as natural cubic spline with five knots based on percentiles). ^3^Vaccinated defined as ≥14 days after receipt of the second dose of any mRNA vaccine. ^4^Defined based on evidence from enzyme-linked immunosorbent assays (ELISA) targeting SARS-CoV-2 receptor-binding domain and spike protein conducted at the Influenza Research Institute at University of Wisconsin-Madison on serum samples collected before enrollment, molecular SARS-CoV-2 test results from clinic testing before enrollment, and self-report at study enrollment. ^5^Delta variant identified in >50% of samples sequenced in Wisconsin after June 21, 2021; 97% of 29 viruses sequenced from samples collected from study participants between June 21, 2021-October 7, 2021 were Delta variant. Participants with SARS-CoV-2 infection between November 3, 2020-June 20, 2021 were excluded.

## Discussion

In this prospective rural community cohort with active illness surveillance, mRNA vaccines were 56% effective against symptomatic and asymptomatic SARS-CoV-2 infection. VE estimates were lower than reported estimates from clinical trials and observational studies based on clinical testing conducted soon after vaccines became available [6, 8, 9].

Our findings of lower VE against SARS-CoV-2 infection during a period with increased circulation of Delta is consistent with previous studies among healthcare workers, nursing home residents, and in retrospective, population-based cohort studies [2-5, 10]. Waning of vaccine protection is also possible, as lower VE was observed with longer time since vaccination in several studies [2, 3, 11]. However, assessment of the contribution of waning immunity and the Delta variant to the observed reduced vaccine protection is complicated by the local increase in Delta circulation coinciding with the period when most cohort members were >5 months from receipt of their second dose. Further studies with additional follow-up time after vaccination for all ages are needed to better understand the impact of waning protection and the importance of booster doses.

This study had several limitations. Relatively few cases occurred during the follow-up period with most vaccinated cases occurring when Delta predominated. The small sample size led to wide confidence intervals and limited our ability to control for potential confounding factors in VE estimates such as preexisting conditions, occupation, and behaviors, which may be associated with vaccination status, vaccine product received, and infection risk. Finally, the study population is largely non-Hispanic White and from a single rural community in central Wisconsin so findings may not be generalizable to other rural communities or other racial and ethnic groups.

Strengths of this study include active follow-up of participants for new illness that included weekly respiratory samples collection for SARS-CoV-2 testing for half of the participants during most of the follow-up period. Weekly surveillance combined with clinical SARS-CoV-2 test results available from linked health records allowed comprehensive capture of SARS-CoV-2 infections. Second, MCHS’s data exchange with the Wisconsin Immunization Registry allowed more accurate classification of vaccination status over time and product received. Third, our analysis included adolescents and rural community members, who have been underrepresented to date. Finally, prior SARS-CoV-2 infections were captured by self-report and serologic testing, reducing the potential for biased VE estimates.

This study demonstrates that two doses of mRNA vaccine reduce the risk of SARS-CoV-2 infection. However, vaccinated persons continue to be at risk for infection in the community, serving as a reminder of the importance of layered prevention measures to break chains of transmission. A booster dose may help increase protection among vaccinated persons, but efforts to increase uptake are essential. Increasing uptake of the primary vaccine series in rural areas, where there is greater hesitancy to receive COVID-19 vaccine and the burden of COVID-19 and associated mortality has been higher than in urban areas, should be a priority [12, 13]. As SARS-CoV-2 continues to circulate and evolve and the COVID-19 vaccination program matures, continued monitoring of COVID-19 VE in the general population is needed.

## Data Availability

Data produced in the present study may be available upon reasonable request to the authors.

## Disclaimer

The findings and conclusions in this report are those of the authors and do not necessarily represent the official position of the US Centers for Disease Control and Prevention.

## Acknowledgements

We thank the following for their contributions to the study: Roxy Eibergen, Lynn Ivacic, Diane Kohnhorst, Erik Kronholm, DeeAnn Polacek, Carla Rottscheit, Elizabeth Armagost, Bobbi Bradley, Hannah Berger, Adam Bissonnette, Joshua Blake, Thomas G. Boyce, Keegan Brighton, Gina Burbey, Deanna Cole, Leila L. Deering, Cody DeHamer, Rachel Fernandez, Sherri Guzinski, Kayla Hanson, Erin Higdon, Jacob P. Johnston, Julie M. Karl, Taylor Kent, Burney A. Kieke Jr., Sarah Kohn, Sarah L. Kopitzke, Tamara Kronenwetter Koepel, Stacey Kyle, Eric LaRose, Kate Lassa, Carrie Marcis, Karen McGreevey, Nidhi Mehta, Daniel Miesbauer, Jennifer Moran, Pamela Mundt, Lisa Ott, Nan Pan, Cory Pike, Rebecca Pilsner, Martha Presson, Nicole Price, Mark Riley, Jacklyn L. E. Salzwedel, Juan Saucedo, Kristin Seyfert, Alex Slenczka, Elisha Stefanski, Robert Strenn, Sandra K. Strey, Melissa Strupp, and Krishna C Upadhyay at Marshfield Clinic Research Institute; Lizheng Guan and Peter Jester at University of Wisconsin-Madison; Fatimah Dawood, Constance Ogokeh, Carrie Reed, and Douglas Slaughter at Centers for Disease Control and Prevention.

## Supplemental Materials

### Supplemental Methods

#### Marshfield Epidemiologic Study Area (MESA)

The Central region of MESA is a geographically-defined population cohort of approximately 53,000 residents residing in 14 zip codes that includes Marshfield, Wisconsin and surrounding area [1]. In this area, 89% of residents receive most of their care from Marshfield Clinic Health System (MCHS), and the population size is similar to US census estimates for the MESA zip codes [1]. The MESA population is predominantly non-Hispanic White, and the population is older and more rural compared to the entire state.

#### Cohort eligibility criteria

For this prospective study, we sampled and recruited MESA residents in strata defined by age group. Young children (age <10 years) and older adults (age ≥70 years) were oversampled. Since enrollment was lower than expected, we opened enrollment to eligible individuals who were not sampled beginning January 2021. We excluded residents of institutional or group settings, individuals who planned to move or reside outside of the study area for ≥21 continuous days during the next 12 months, and participants in COVID-19 clinical trials. No exclusions were made for individuals who had had SARS-CoV-2 infection prior to enrollment, nor for individuals who had received COVID-19 vaccine doses prior to enrollment.

#### Surveillance group assignment

Frequency of respiratory sample collection, weekly or with qualifying illness, was initially randomly assigned in 1:1 ratio from initiation of enrollment through December 28, 2020. However, because of slower than expected enrollment and concurrent increased SARS-CoV-2 circulation locally, enrollments from December 29, 2020 through January 25, 2021 were assigned to weekly respiratory sample collection. Enrollments after January 25, 2021 were assigned to respiratory sample collection with qualifying illness except children aged <10 and Spanish-speaking participants who were assigned to weekly respiratory sample collection until enrollment targets were reached, because of low initial participation in these groups.

#### Data and serum sample collection

At enrollment, consenting participants completed a survey on demographics, current health status, medical history, prior SARS-CoV-2 infection and exposure history, and relevant environmental and behavioral risk factors. Serum samples were collected at enrollment and approximately 12 and 24 weeks later. Additional information regarding demographics, SARS-CoV-2 clinical test dates and results, COVID-19 and influenza vaccinations, preexisting conditions, and healthcare visits were extracted from MCHS’s electronic health records. MCHS exchanges vaccination data with the Wisconsin Immunization Registry weekly. If self-report of vaccination was not documented in the electronic health records, vaccination dates, vaccine product, and location of vaccination were obtained from the participant’s COVID-19 vaccination card.

#### COVID-19 vaccine roll-out in Wisconsin

In Wisconsin, the initial phases of COVID-19 vaccine roll-out began December 16, 2020 and prioritized vaccination of frontline healthcare workers, first responders, residents of long-term care facilities, and persons aged ≥65 years. Vaccine became available on March 1, 2021 to those enrolled in Medicaid long-term care programs, those who work or live in congregate living facilities, public-facing essential workers, and non-frontline essential healthcare personnel. Individuals aged ≥16 years with certain medical conditions that have greater risk of severe infection and all individuals aged ≥16 years were eligible March 22 and April 5, 2021, respectively. Adolescents aged 12-15 years were vaccine eligible on May 13, 2021.

#### Laboratory methods

Respiratory samples collected during cohort surveillance were tested by real-time reverse transcription polymerase chain reaction (rRT-PCR) for SARS-CoV-2 using the ThermoFisher Combo Kit platform at Marshfield Clinic Research Institute’s Integrated Research and Development Laboratory [2]. A subset of positive samples (e.g., samples with cycle threshold values <30) collected between April 12, 2021-October 7, 2021 were sequenced using a modified ARTIC sequencing methodology (https://artic.network/ncov-2019). A laboratory-confirmed SARS-CoV-2 infection was defined in a participant who had a specimen collected during surveillance that was positive by rRT-PCR or a specimen collected for clinical purposes that was tested by a PCR-based assay conducted at MCHS.

Serum samples collected at the time of enrollment were tested for SARS-CoV-2 antibodies using an enzyme-linked immunosorbent assay (ELISA) that targeted the SARS-CoV-2 receptor-binding domain, the full-length spike (S1S2) protein, and nucleocapsid protein following standard procedures at the Influenza Research Institute at University of Wisconsin-Madison [3]. Specimens collected from participants who were unvaccinated at the time of enrollment were classified as seropositive based on a Bayesian hierarchical model considering reactivity against S1S2 and the receptor-binding domain targets.

---

**Supplemental Table.** Characteristics of the study cohort

|  |  | Vaccination status<sup>1</sup> |  |  |  |
| --- | --- | --- | --- | --- | --- |
|  | All<br>(n=1266) | Received<br>Moderna<br>COVID-19<br>vaccine<br>(n=329) | Received<br>Pfizer-BioNTech<br>COVID-19<br>vaccine<br>(n=608) | Not<br>vaccinated<br>(n=329) | SARS-CoV-2<br>infection<sup>2</sup><br>(n=118) |
| Age group<sup>3</sup> |  |  |  |  |  |
| 12-17 years | 129 (10) | 0 | 84 (14) | 45 (14) | 11 (9) |
| 18-49 years | 470 (37) | 91 (28) | 219 (36) | 160 (49) | 58 (49) |
| 50-64 years | 272 (21) | 76 (23) | 133 (22) | 63 (19) | 23 (19) |
| ≥65 years | 395 (31) | 162 (49) | 172 (28) | 61 (19) | 26 (22) |
| Sex |  |  |  |  |  |
| Female | 747 (59) | 188 (57) | 383 (63) | 176 (54) | 70 (59) |
| Male | 519 (41) | 141 (43) | 225 (37) | 153 (46) | 48 (41) |
| Race/ethnicity<sup>4</sup> |  |  |  |  |  |
| Non-Hispanic White | 1203 (95) | 321 (98) | 576 (95) | 306 (93) | 111 (94) |
| Hispanic | 39 (3) | 4 (1) | 18 (3) | 17 (5) | 5 (4) |
| Non-Hispanic non-White | 20 (2) | 3 (1) | 12 (2) | 5 (2) | 2 (2) |
| Preexisting medical<br>condition<sup>5</sup> |  |  |  |  |  |
| No | 547 (43) | 110 (33) | 281 (46) | 156 (47) | 55 (47) |
| Yes | 719 (57) | 219 (67) | 327 (54) | 173 (53) | 63 (53) |
| Asthma | 109 (9) | 27 (8) | 46 (8) | 36 (11) | 6 (5) |
| Cancer | 31 (2) | 15 (5) | 10 (2) | 6 (2) | 3 (3) |
| Chronic kidney disease | 26 (2) | 7 (2) | 13 (2) | 6 (2) | 1 (1) |
| COPD | 31 (2) | 11 (3) | 13 (2) | 7 (2) | 3 (3) |
| Hypertension | 307 (24) | 115 (35) | 142 (23) | 50 (15) | 17 (14) |
| Immunocompromised | 47 (4) | 15 (5) | 20 (3) | 12 (4) | 5 (4) |
| Serious heart condition | 72 (6) | 34 (10) | 24 (4) | 14 (4) | 5 (4) |
| Type 2 diabetes | 108 (9) | 41 (12) | 48 (8) | 19 (6) | 9 (8) |
| Obese | 471 (37) | 136 (41) | 208 (34) | 127 (39) | 51 (43) |
| Receipt of 2020-2021<br>influenza vaccine |  |  |  |  |  |
| No | 382 (30) | 63 (19) | 125 (21) | 194 (59) | 46 (39) |
| Yes | 884 (70) | 266 (81) | 483 (79) | 135 (41) | 72 (61) |
| Health insurance type<sup>4</sup> |  |  |  |  |  |
| Private | 804 (64) | 169 (51) | 425 (70) | 210 (64) | 83 (71) |
| Public | 435 (34) | 156 (48) | 173 (29) | 106 (33) | 31 (27) |
| None | 21 (2) | 4 (1) | 7 (1) | 10 (3) | 3 (3) |
| Occupation/Industry<sup>6</sup> |  |  |  |  |  |
| Healthcare/Social services | 241 (36) | 45 (30) | 166 (51) | 30 (16) | 20 (25) |
|  | All<br>(n=1266) | Received<br>Moderna<br>COVID-19<br>vaccine<br>(n=329) | Received<br>Pfizer-BioNTech<br>COVID-19<br>vaccine<br>(n=608) | Not<br>vaccinated<br>(n=329) | SARS-CoV-2<br>infection<sup>2</sup><br>(n=118) |
| Manufacturing | 78 (12) | 15 (10) | 25 (8) | 38 (20) | 7 (9) |
| Education | 75 (11) | 33 (22) | 23 (7) | 19 (10) | 13 (16) |
| Retail | 49 (7) | 5 (3) | 22 (7) | 22 (12) | 6 (8) |
| Other service | 38 (6) | 8 (5) | 21 (6) | 9 (5) | 3 (4) |
| Other industry | 186 (28) | 46 (30) | 68 (21) | 72 (38) | 30 (38) |
| Frequency of respiratory<br>sample collection<sup>7</sup> |  |  |  |  |  |
| Weekly swabbing | 635 (50) | 174 (53) | 295 (49) | 166 (50) | 67 (57) |
| Swabbing with qualifying<br>illness | 631 (50) | 155 (47) | 313 (51) | 163 (50) | 51 (43) |
| Prior SARS-CoV-2 infection<sup>8</sup> |  |  |  |  |  |
| No | 1088 (86) | 292 (89) | 523 (86) | 273 (83) | 114 (97) |
| Yes | 178 (14) | 37 (11) | 85 (14) | 56 (17) | 4 (3) |

Data are no. (%) of participants.
Abbreviations: COVID-19, coronavirus disease 2019; SARS-CoV-2, severe acute respiratory syndrome coronavirus 2; COPD, chronic obstructive pulmonary disease
<sup>1</sup>Documented receipt of 2 doses before the time of censoring.
<sup>2</sup>New laboratory confirmed SARS-CoV-2 infections occurring during the follow-up period (November 3, 2020-December 7, 2021) among two-dose vaccinated or unvaccinated participants.
<sup>3</sup>Age as of end of follow-up (or censoring)
<sup>4</sup>Data missing or unknown; % based on those with data
<sup>5</sup>Preexisting medical condition includes self-report of asthma, cancer, chronic kidney disease, COPD, hypertension, immunocompromised state, serious heart condition, and type 2 diabetes, and obesity defined as BMI $\geq 30$ calculated from self-reported height and weight
<sup>6</sup>Percent among employed participants; based on the North American Industry Classification System (NAICS) grouping of 20 large industry sectors (https://www.census.gov/naics/).
<sup>7</sup>Participants assigned to swab weekly for the first 26 weeks regardless of symptoms and with a qualifying illness after study week 27 or to swab with a qualifying illness for the duration of follow-up. A qualifying illness was an illness with new onset in the past 7 days and at least one of the following symptoms: cough, fever or chills, sore throat, muscle or body aches, loss of smell or taste, shortness of breath, or diarrhea.

**Supplemental Figure.**
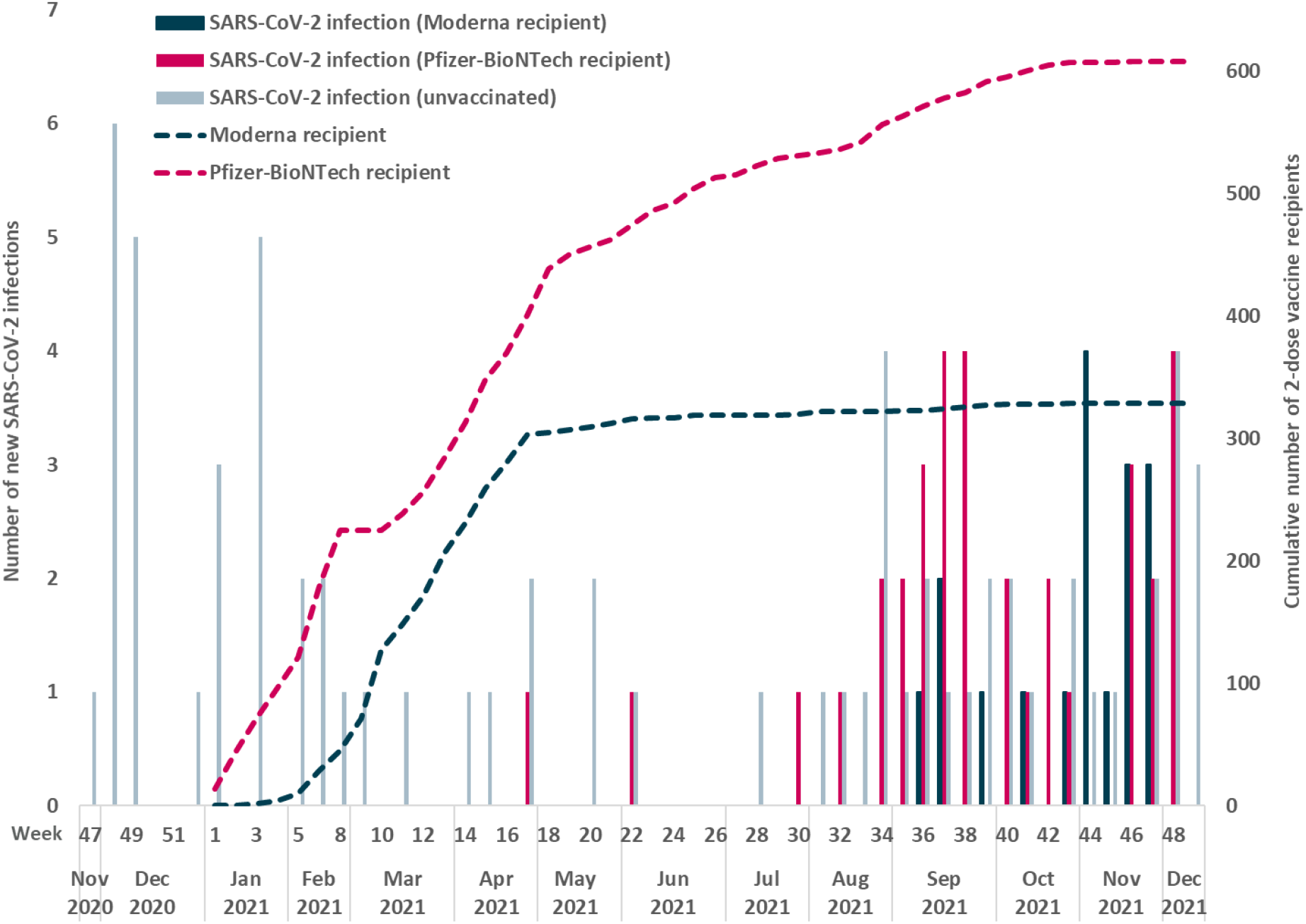
Number of new SARS-CoV-2 infections during the follow-up period (November 3, 2020 through December 7, 2021), and cumulative number of two-dose vaccinated participants by week.

